# Deep learning-based risk stratification of preoperative breast biopsies using digital whole slide images

**DOI:** 10.1101/2023.08.22.23294409

**Authors:** Constance Boissin, Yinxi Wang, Abhinav Sharma, Philippe Weitz, Emelie Karlsson, Stephanie Robertson, Johan Hartman, Mattias Rantalainen

**Affiliations:** Department of Medical Epidemiology and Biostatistics, Karolinska Institutet, 17177 Stockholm, Sweden; Department of Oncology and Pathology, Karolinska Institutet, 17176 Stockholm, Sweden; Department of Clinical Pathology and Cytology, Karolinska University Laboratory, 11883 Stockholm, Sweden; MedTechLabs, BioClinicum, Karolinska University Hospital, 17176 Stockholm, Sweden

**Author notes:** Corresponding author: Mattias Rantalainen.

## Abstract

**Introduction:** Nottingham histological grade (NHG) is a well established prognostic factor in breast cancer histopathology. However, manual NHG assessment of biopsies is challenging and has a large inter-assessor variability with a large proportion being classified as NHG2 (intermediate grade). Here, we evaluate whether DeepGrade, a previously developed model for the risk stratification of resected tumour specimens, could be applied to risk-stratify biopsy specimens.

**Methods:** A total of 11,943,905 tiles from 1171 whole slide images (WSIs) of preoperative biopsies from 897 patients diagnosed with breast cancer in Stockholm, Sweden, were included in this retrospective observational study. DeepGrade, a deep convolutional neural network model, was applied for classification of low and high risk tumours and evaluated against clinically assigned grades 1 and 3 using area under the operating curve (AUC). The prognostic value of the DeepGrade model in the biopsy setting was evaluated using time-to-event analysis.

**Results:** The DeepGrade model classified resected tumour cases with grades NHG1 and NHG3 using only biopsy specimens with an AUC of 0.903 (95% CI: 0.88;0.93). The model could also classify the biopsy NHG (1 and 3) assessed on the biopsy of 186 patients with an AUC of 0.959 (95% CI: 0.93; 0.99). Furthermore, out of the 434 NHG2 tumours, 255 (59%) were classified as DeepGrade2-low, and 179 (41%) were classified as DeepGrade2-high. Using a multivariable Cox proportional hazards model the hazard ratio between low- and high-risk groups was estimated as 2.01 (p-value = 0.036).

**Conclusions:** DeepGrade could predict the resected tumour grades NHG1 and NHG3 using only the biopsy specimen and sub-classify grade 2 tumours into low and high risks. The results demonstrate that the DeepGrade model can provide decision support for biopsy grading, and potentially provide decision support in the clinical setting to identifying high-risk tumours based on preoperative breast biopsies, thus improving information available for clinical treatment decisions.

## Introduction

Breast cancer is now the most common cancer type globally^1^. In the majority of cases, suspected breast lesions are initially identified by mammography screening which is recommended in most developed countries to prevent breast cancer^2,3^. For women with a suspected lesion, a preoperative core biopsy is performed for histological inspection of the tissue^4^. Evaluation of the biopsy by a pathologist is key to diagnose breast cancer, but also for treatment decisions based on histological subtypes and biomarker analysis^5^.

In addition to histopathological diagnosis and subtyping, grading is a cornerstone in the histopathological assessment of the biopsy^6^. Histological grade reflects the degree of differentiation of a tumour by comparing the similarity of malignant cells to that of normal breast terminal duct lobular units^7^. Currently, the most commonly used grading method is the Notthingham Histological Grade (NHG) adapted by Elston-Ellis following work from Bloom-Richardson^8,9^. Histological grading relies on the performance and expertise of pathologists which evaluate three morphological features: the degree of tubular formation (gland architecture), nuclear pleomorphism (nucleus size and shape) and the mitotic count^9^. Histological grade is also an important prognostic feature, with higher grade tumours being associated with poor prognosis independently of the tumour type or endocrine status^10^. Therefore, it has an important role in deciding what type of treatment a patient will receive^11^. One of the major challenges with histological grading is that it relies on the experience, expertise and interpretation of the pathologist, resulting in high inter-observer and inter-lab variabilities^7,12^. In addition, about 50% of all resected breast cancer specimens are diagnosed as NHG2, which has no clinical value^12–14^.

Histological grade assessment is even more complicated in biopsy specimens with very limited tumour material and frequent tissue artefacts^12^. This causes significant discrepancies between the biopsy grade, and the histological grade assigned on the resected specimen^15^. These uncertainties also come with a greater number of biopsy samples not being assigned a grade at all, and with up to 70% of the biopsy samples being assigned the intermediate NHG2 grade^13,16,17^.

The recent advances in computational pathology based on the availability of large amounts of digitised whole-slide histopathological images (WSIs), as well as the development of novel artificial intelligence technologies, has enabled model-based grading of tumours in resected specimens, and also enabled further risk-stratification of intermediate-risk NHG2 patients into two risk subgroups with independent prognostic value^18^.

In this study, we aim to assess whether the DeepGrade model, developed using resected tumour specimens, could be used to predict NHG1 and NHG3 tumours in the biopsy specimens. It will allow the earlier identification of the high risk tumours from the initial biopsy specimens, provide decision support for grading of preoperative biopsies, and further improve information that can be used in the treatment planning at the preoperative stage.

## Methods

### Patients

This retrospective study included female patients who underwent a biopsy at the Stockholm South General Hospital in Stockholm, Sweden between June 2012 and May 2018. Patients diagnosed with invasive breast cancer as their primary diagnosis and who had undergone a surgical removal of their tumour within two months following their biopsy without receiving neoadjuvant therapy were included in the study (see Figure 1 for detailed explanation of selection criteria). A total of 1171 whole slide images (WSIs) from 897 patients were included in the final analyses. The WSIs from the resected specimen of 807 of these patients was also available which were also used for comparison of the prediction of DeepGrade score on this material. Clinical data was retrieved retrospectively, either from the Swedish National Breast Cancer Registry, or from the pathology reports. This study was reviewed and approved by local and governmental ethics committees.

**Figure 1.**
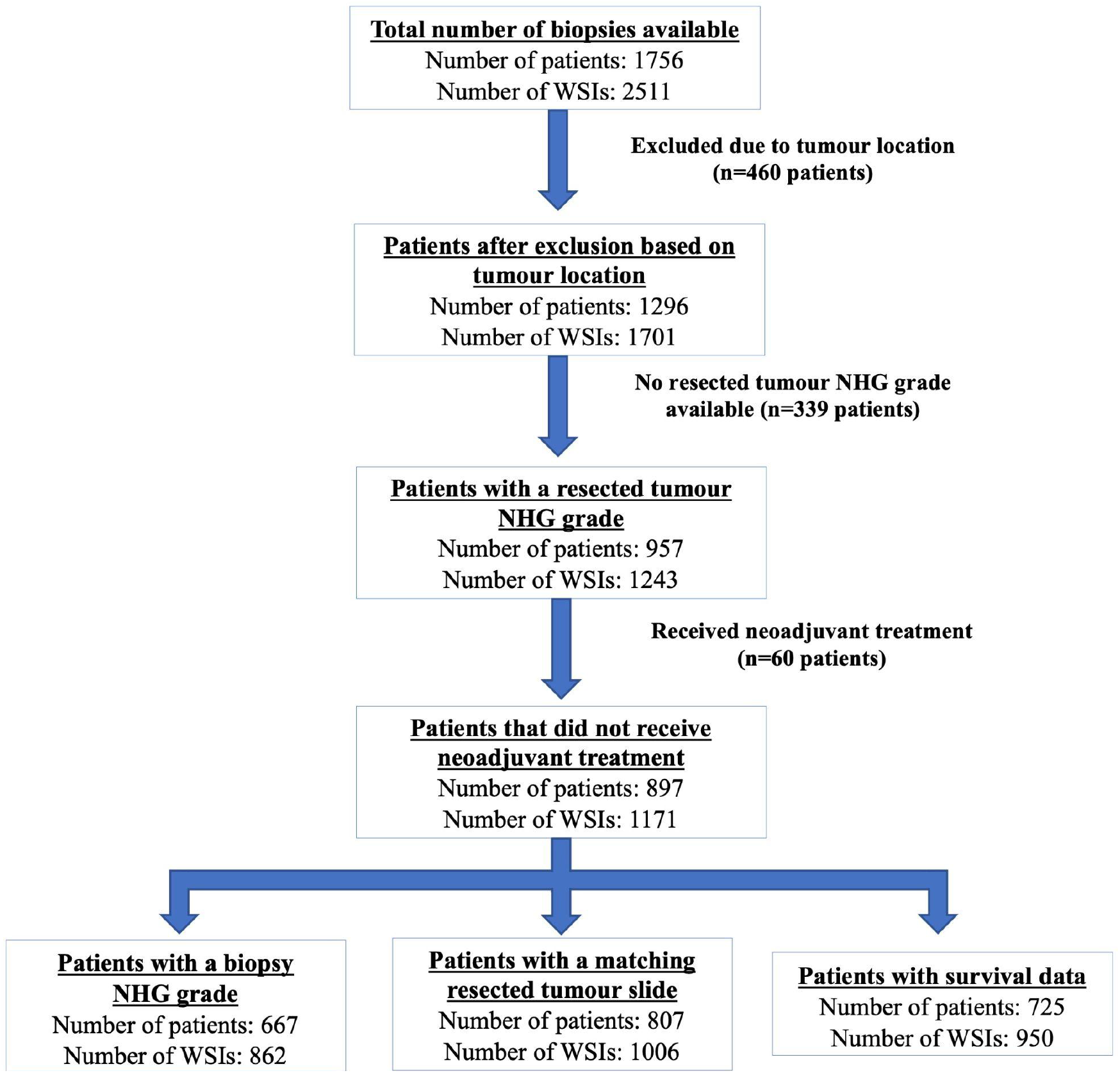
Consort diagram. The data used contained whole slide images of biopsies for 897 patients who have a resected tumour NHG grade and who did not receive neoadjuvant chemotherapy. A total of 667 patients had a biopsy NHG grade, 807 patients had a matching resected tumour slide on which DeepGrade predictions could also be performed and, survival data was available for 726 patients.

### WSI and deep model

For each patient between one and seven Haematoxylin and Eosin (H&E) stained formalin-fixed paraffin-embedded (FFPE) histopathology slides of biopsy specimens were digitised in-house using either Hamamatsu Nanozoomer XR or Hamamatsu Nanozoomer S360 scanners at 40X magnification (0.227 μm/pixel and 0.230 μm/pixel respectively). Methodology for preprocessing of the WSIs was performed according to the methodology previously described^18^. Initially, tissue segmentation was performed by transforming lower-level representations obtained using OpenSlide^19^ from RGB to HSV colour space. Two masks were then generated using the Otsu’s threshold to remove the non-tissue areas^20^. A maximum value of 25 was added to the Otsu threshold value in order to reduce the removal of the tissue regions due to the high threshold value on the transformed saturation channel in some cases.

WSI regions included in the tissue mask were tiled into image tiles of 598×598 pixels with a down-sampled resolution equivalent to 20X (271μm × 271μm). Due to the small tissue area in biopsy specimens, tiling was performed with 75% overlap between two consecutive tiles. Next, in order to remove unsharp tiles, any remaining tiles with background, and those with adipose tissue, blurred tiles were excluded when a variance of the Laplacian filtering was lower than 500^21^. Lastly, to address the stain variabilities in WSIs, colour normalisation across each WSI was performed using the method described by Macenko et al.^22^, and as implemented by Wang et al^18^. For 807 patients with preoperative biopsies with a matching resected tumour WSI, a similar pre-processing method was performed for the WSI pre-processing with two significant changes. First, no overlap between two consecutive tiles was considered. Secondly, after the colour normalisation step, a tumour segmentation model developed^18^ was applied to include only the tiles from the invasive cancer regions in the resected specimens for further downstream analysis. After preprocessing, a total of 11,943,905 tiles were used for predictions from biopsy specimens, and 1,169,316 were used from the surgical specimens.

### Histological grade prediction

Classification of NHG3 versus NHG1 on the biopsy WSI, was performed using an ensemble of 10 CNN models that were chosen randomly out of the ensemble of 20 models that were developed as the DeepGrade model^18^. All models were developed using an Inception V3 architecture^23^. The initial models were trained on 844 WSIs, of which 166 patients for whom the biopsy WSI were also included in this study. DeepGrade models were trained to classify NHG3 and NHG1 tumours in WSIs from resected specimens. Each model in DeepGrade outputs the two class prediction probabilities for each tile (P(NHG_3_|tile_i_) and P(NHG_1_|tile_i_)). The P(NHG_3_|tile_i_) class probability from each of the ten models in the ensemble were averaged to provide the tile-level prediction score. Next, for each patient all the tile-level prediction scores of all the WSIs were pooled together and the patient-level prediction score was aggregated by considering the upper-percentile (99th) of the pooled tile-level prediction scores. Binary classification threshold into low and high risk groups was determined using the Youden’s J statistic^24^ and was calculated separately for the biopsy and the surgical WSIs. For NHG1 and NHG3 DeepGrade-low represents NHG1 whereas DeepGrade-high represents NHG3. For NHG2 tumours, DeepGrade stratification represents risk groups which are NHG1-like or NHG3-like. For NHG1 and NHG3, prediction performance of the DeepGrade model was evaluated against clinically assigned NHG grade on both the biopsy specimen (biopsy NHG grade) and on the surgically resected specimen (resected tumour NHG grade) by a pathologist. The classification performances were measured using the receiver operating characteristic (ROC) curves and the linked area under the curve (AUC) using R package pROC^25^. Agreement between the assigned NHG grade (from the biopsy specimen or the resected tumour specimen) and the obtained DeepGrade was measured using Cohen’s kappa and the following interpretations: 0-0.20: slight agreement, 0.21-0.40: fair agreement, 0.41- 0.60: moderate agreement, 0.61-0.80: substantial agreement and 0.81-1.00: almost perfect agreement^26,27^. The classification performance of the DeepGrade on the biopsy WSI was also compared to the classification performed on the surgical WSI. Finally, the rates of recurrence-free survival (RFS) as defined by the presence of a locoregional or distant metastasis or death were compared between patients who were assigned in the DeepGrade-high and DeepGrade-low groups. The time-to-event was defined as the number of days between the date of initial diagnosis and either date of recurrence or loss of follow-up. The R packages ‘survival’ and ‘survminer’ were used to visualise the survival outcomes between groups, and the ‘forestmodel’ package was used to estimate adjusted hazard ratios (HRs) using multivariate Cox proportional hazards regression models.

## Results

Firstly, we assessed the discrepancies between the clinical assignment of NHG grade on the biopsy and resected specimens (Figure 2a). A quarter of the patients did not have NHG grade assigned on the biopsy specimen at all in clinical routine, and 72% of the patients who had a biopsy NHG grade available were of NHG2 grade. The overall agreement between the clinical grade assignments on the biopsy and on the resected tumour specimen was 65.4% when including patients for whom we have both diagnoses. We observed a fair agreement with the Cohen’s kappa value of 0.40 (95% CIs: 0.34;0.46) between them.

**Figure 2.**
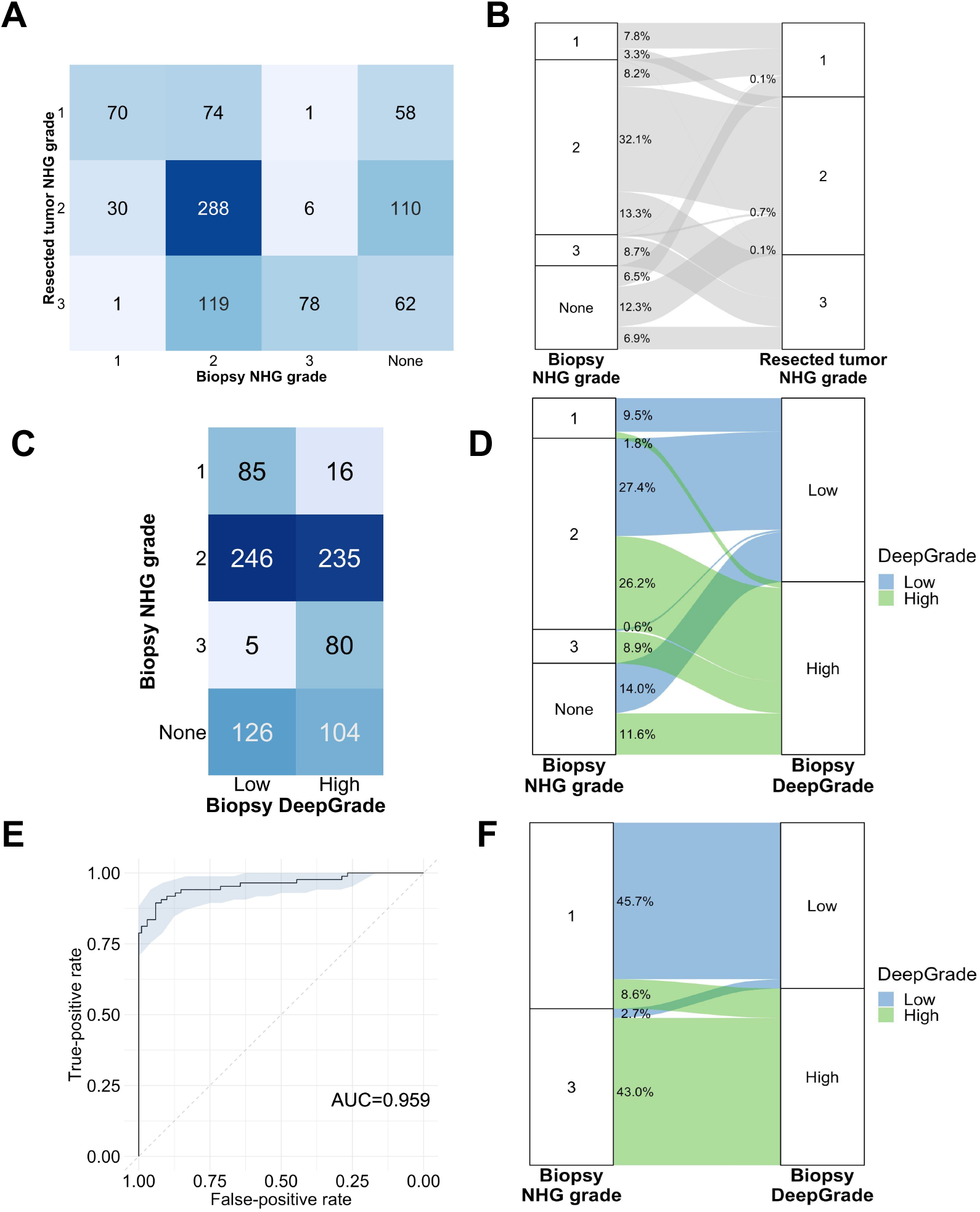
DeepGrade prediction results obtained on biopsy specimens compared to the clinical NHG assigned by the pathologist. Comparison between NHG grades assigned by pathologists on the biopsy specimen and the resected tumour specimen; A. confusion matrix, B. Sankey plot. Comparison between NHG grades assigned by pathologists on the biopsy specimen and the DeepGrade assigned on the biopsy specimen; C. confusion matrix, D. Sankey plot. E. ROC curve of the prediction score obtained by the DeepGrade CNN model compared to biopsy NHG grades 1 and 3. F. Sankey plot representing the number of patients assigned to each of the Deep Grade risk group by biopsy NHG grades 1 and 3.

### Assessment of the DeepGrade classification performance on the biopsy specimen

We evaluated the risk classification performance of the DeepGrade model on the NHG1 and NHG3 grade biopsy specimens. We observed an AUC score of 0.959 (95% CI: 0.930;0.989), see Figure 2E. For 165 out of 186 patients (89.30%) the DeepGrade model and the pathologists were in agreement, representing a substantial agreement with a kappa value of 0.77.

Of the 897 patients, only 0.6% (5) who had a biopsy NHG3 grade were assigned to the DeepGrade low-risk group, which corresponds to NHG1 (Figure 2D). Out of the 230 patients who were not assigned a NHG grade at biopsy, 126 (54.8%) were classified in the DeepGrade low-risk group, and 104 (45.2%) were classified in the high-risk group.

### Assessment of the DeepGrade classification performance compared to the resected tumour specimen

To assess the hypothesis that the DeepGrade model not only to predict the NHG grade of the biopsy, but also predict the clinically assigned NHG1 and NHG3 grades assigned on the resected specimens using only the biopsy material, we compared the prediction results obtained. We observed an AUC score of 0.903 (95% CI: 0.876;0.930), see Figure 3A. The agreement between the predictions obtained and that of the pathologist was of 385 out of 463 patients (83.15%) and the kappa value was 0.66 meaning substantial agreement. When looking at the patients who were in the DeepGrade-low risk group, but who had a resected tumour NHG3 grade, the biopsy NHG grade was either NHG2 or no NHG grade in 93% of the cases, with only 3 patients who had NHG3 grade at both biopsy and resected tumour level. Out of the 434 patients who had a resected tumour with NHG2 grade, 259 (59.67%) were assigned to the DeepGrade low-risk group while 179 (40.32%) were assigned to the DeepGrade high-risk group.

**Figure 3.**
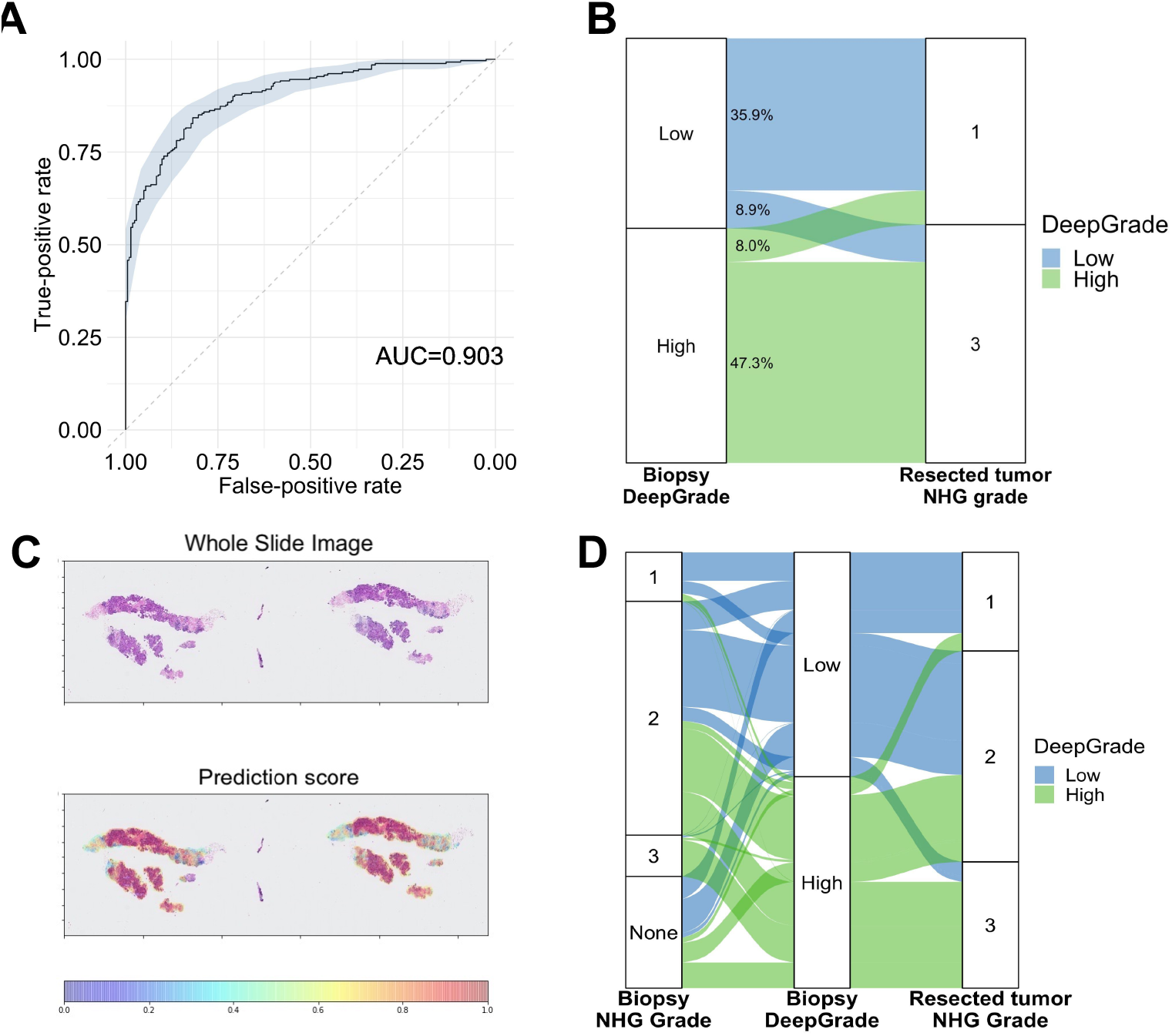
DeepGrade prediction results obtained on biopsy specimens compared to the clinical NHG assigned by the pathologist on the resected specimen. A. ROC of the continuous prediction score versus the resected tumour NHG grades 1 and 3 assigned by a pathologist. B. Sankey plot of the proportion of patients predicted with Deep Grade High and Low versus the resected tumour NHG grade. C. Example of a whole slide image with prediction results. Red is more likely to be predicted as High risk. D. Sankey plot with results of all biopsy specimen comparing the obtained Deep Grade with both the biopsy NHG grade and the resected tumour NHG grade

### Comparison between DeepGrade score on biopsy and resected tumour specimens

To verify whether the results obtained on the biopsy specimens are in line with those obtained on the resected tumour specimen, we compared the DeepGrade score for 807 patients for which we had both specimens available (Figure 4). Almost three quarters of the patients were assigned the same DeepGrade score on the biopsy and resected specimens. This proportion is even higher when considering only patients with a resected tumour NHG1 or NHG3, with 81.3% of the patients who were assigned the same DeepGrade score.

**Figure 4.**
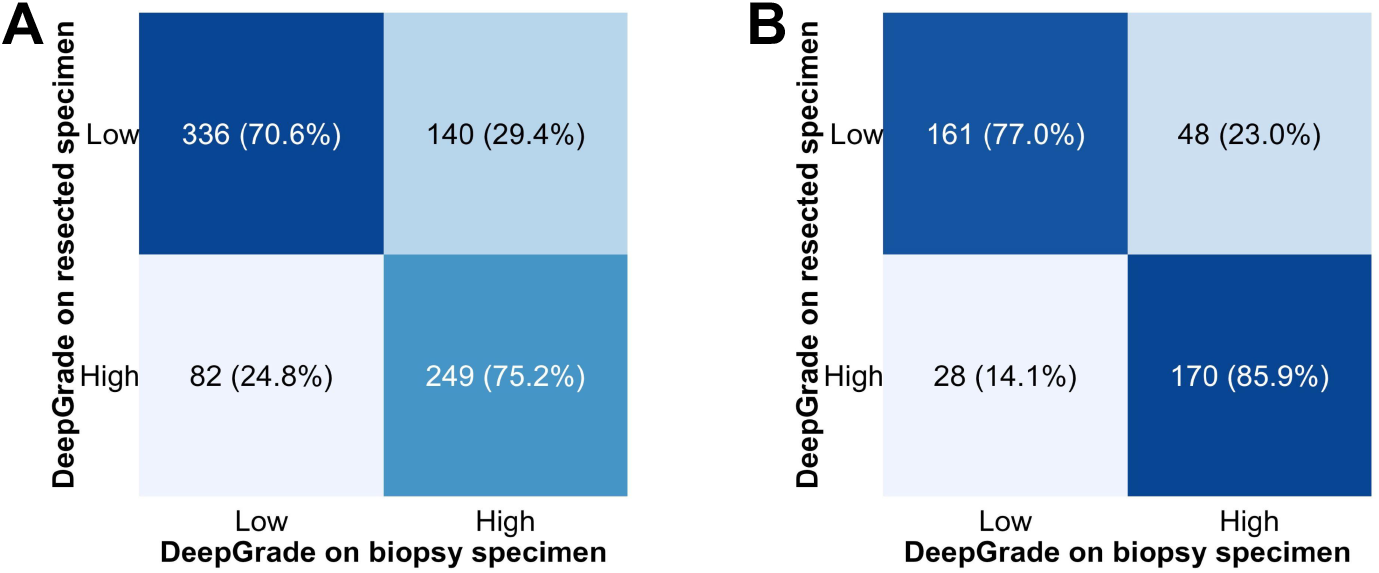
Comparison of DeepGrade scores on biopsy and resected specimens. A. Confusion matrix for all NHG grades combined. B. Confusion matrix for only resected NHG1 and NHG3 grades.

### Prognostic performance of the DeepGrade score

The prognostic performance of the DeepGrade model in biopsy specimens was measured based on recurrence-free survival and was visualised using Kaplan-Meier curves. The independent prognostic value was measured using multivariable Cox Proportional Hazards model adjusting for age (resembling information available at the biopsy stage). When including all patients, the DeepGrade model was found to be a predictor of survival with an estimated hazard ratio of 2.01 for patients with DeepGrade-high on biopsy specimen compared to those with a DeepGrade-low score, independently of the patient’s age (Figure 5).

**Figure 5.**
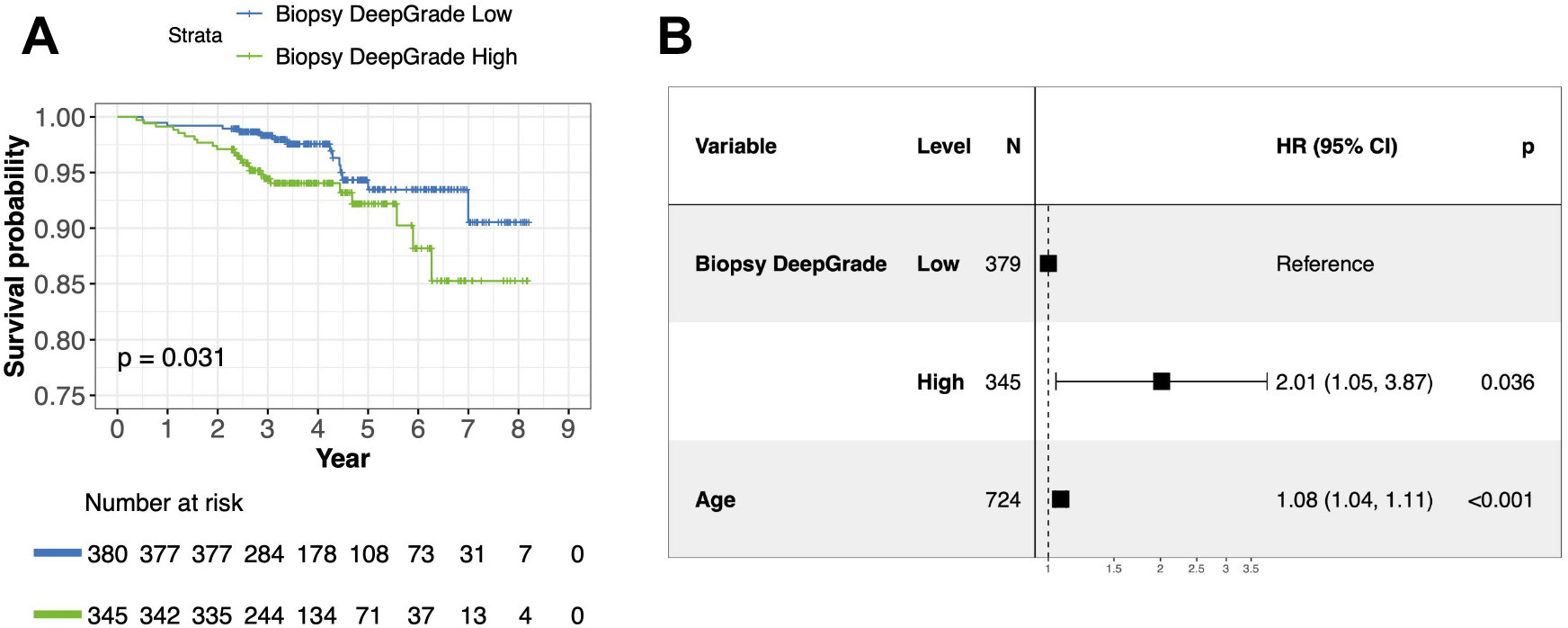
Recurrence-free survival outcomes for breast cancer patients by DeepGrade score obtained on the biopsy specimen. A. Kaplan-Meier curves for patients stratified by biopsy DeepGrade low and high risk groups. High-risk group had the worst prognosis. B. Forest plot from multivariable Cox proportional hasard regression including the biopsy DeepGrade score and age at diagnosis.

## Discussion

The aim of this study was to investigate whether the DeepGrade model, previously developed to risk-stratify patients based on the resected tumour specimens, could also be used to risk-stratify biopsy specimens. We observed a high classification performance when comparing the DeepGrade predictions on the biopsy specimen to the biopsy NHG grade. Most interestingly, the DeepGrade model could predict the grade of the resected tumour specimen while analysing only biopsy material and classification of patients using the DeepGrade model was predictive of survival at time of biopsy.

The identification of NHG3 grade patients at the time of biopsy is critical for the decision to treat a patient with neoadjuvant chemotherapy^7,28^, however, conventional NHG grading of biopsies remains challenging. Neoadjuvant therapy is now recommended to most HER2+ and triple negative cancers, of whom the vast majority are NHG3. Within the larger ER+, HER2-subgroup there are patients with high-risk tumours that should be considered for neoadjuvant chemotherapy but most biopsies are assigned a NHG2 grade, or are not graded at all^12,16^. This leads to a discrepancy between pathologists, with up to 45% of women who had a change in diagnosis in one cohort^16^. In a similar manner, 46% of patients who were not assigned a grade at the biopsy level were assigned to the high-risk group, of which 50% were actually assigned a NHG3 grade on their resected tumour specimen.

Several studies have developed models to predict grade using deep-learning models on whole-slide images from resected tumour specimens but not core biopsies^18,29–31^. In particular, Wang et al. obtained an AUC of 0.907 in their external data which is in line with the accuracy we obtain in the biopsy specimen when comparing to the resected tumour grades NHG1 versus NHG3 (0.903)^18^. Others who have predicted grade into two groups (low-grade and high-grade) on resected tumour specimens obtained agreements around 80%, and kappa values between 0.59 and 0.64^29,30^. Despite predicting the resected specimen grade using only biopsy material, we get even higher performance results between grades NHG1 and NHG3 with an agreement of 83% and a kappa value of 0.66.

The use of biopsy specimens in computational pathology for breast cancer is relatively rare in the literature, as opposed to the work performed in prostate cancer^32–34^. Besides from the studies mentioned above for tumour identification^35–37^, studies aimed to predict the response to neoadjuvant therapy, in part using grade as their training material^38,39^. The proposed methodology in this study could be used as a decision support tool to complement human pathologists, as it establishes a risk-assessment of all tumours, including those that are hard to grade, and without any pre-established criteria for selection, such as cancer subtype.

This is the first study which suggests to sub-classify grade 2 tumours already at time of biopsy. Although several methods have previously been suggested to sub-classify patients, most use gene expression profiling assays which are time-consuming and remain costly^40^. The sub-classification method presented in this study has the advantages of providing a result to the pathologist in a short time-frame and at a very low-cost given most pathology laboratories in high-income countries already use digitised WSIs^18^. Some of the study limitations which are worth mentioning include the fact that it is based on retrospective material in order to obtain a large enough sample when only including one hospital. Furthermore, for 166 patients (18.5%), their resected specimen was used as training data in the initial Deep Grade model^18^. Finally, this model was evaluated on the whole biopsy tissue area, without prior tumour detection model, it is therefore not impossible that the results are on the lower performance side and that they could achieve even better accuracy in the future if that is to be performed first.

In conclusion, we found that the resected tumour grade could be diagnosed based-on using only biopsy specimens, indicating that high-risk tumours could be identified at the preoperative stage. In the future, this could provide decision support to pathologists as well as treating physicians to improve the quality of relevant information for clinical decisions earlier on in the process, and thus potentially reduce both over- and under-treatment of patients in the neoadjuvant setting.

## Acknowledgments

This project was supported by funding from the Swedish Research Council under the frame of ERA PerMed (ERAPERMED2019-224 - ABCAP), Swedish Research Council, VINNOVA (SwAIPP project), Swedish Cancer Society, Karolinska Institutet (Cancer Research KI; StratCan), MedTechLabs, Swedish e-science Research Centre (SeRC), Stockholm Region, Stockholm Cancer Society and Swedish Breast Cancer Association

## Data availability

The datasets analysed during the current study are not publicly available due to local privacy laws but are available from the corresponding author upon reasonable request.

